# THRomboprophylaxis in Individuals undergoing superficial endoVEnous treatment: a multi-centre, assessor blind, randomised controlled trial – THRIVE trial

**DOI:** 10.1101/2023.12.19.23300215

**Authors:** Matthew Machin, Sarah Whittley, John Norrie, Laura Burgess, Beverley J Hunt, Layla Bolton-Saghdaoui, Joseph Shalhoub, Tamara Everington, Manjit Gohel, Mark Whiteley, Steven Rogers, Sarah Onida, Benedict Turner, Sandip Nandhra, Rebecca Lawton, Annya Stephens-Boal, Carolyn Singer, Joanne Dunbar, Daniel Carradice, Alun Huw Davies

## Abstract

**Introduction:** Endovenous therapy is the first-choice management for symptomatic varicose veins in NICE guidelines, with 56-70,000 procedures performed annually in the UK. Venous thromboembolism (VTE), including deep vein thrombosis (DVT) and pulmonary embolism (PE), and endothermal heat-induced thrombosis (EHIT), are known complications of endovenous therapy, occurring at a rate of up to 3.4%. In an attempt to reduce VTE, 73% of UK practitioners administer pharmacological thromboprophylaxis. However, no high-quality evidence to support this practice exists. Pharmacological thromboprophylaxis may have clinical and cost benefit in preventing VTE, however, further evidence is needed. The aims of this study are to establish whether when endovenous therapy is undertaken: a single dose or course of pharmacological thromboprophylaxis alters the risk of VTE; pharmacological thromboprophylaxis is associated with an increased rate of bleeding events; pharmacological prophylaxis is cost effective.

**Methods and analysis:** A multi-centre, assessor-blind, randomised controlled trial (RCT). We aim to recruit 6660 participants undergoing superficial endovenous interventions under local anaesthesia. Forty sites across the UK, both NHS and private, will be included. Participants will be randomised to either intervention (a single dose or extended course of pharmacological thromboprophylaxis plus compression) or control (compression alone). Participants will undergo a lower limb venous duplex ultrasound scan at 21-28 days post-procedure to identify asymptomatic DVT. The ultrasound duplex scan will be conducted locally by blinded assessors. Participants will also be contacted remotely for follow-up at 7-days and 90-days post-procedure. The primary outcome is imaging confirmed lower limb DVT with or without symptoms, or PE with symptoms within 90 days of treatment. The main analysis will be according to the intention-to-treat principle and will compare the rates of VTE at 90 days, using a repeated measures analysis of variance (ANOVA), adjusting for any pre-specified strongly prognostic baseline covariates using a mixed effects logistic regression.

**Trial registration number:** ISRCTN18501431

**ARTICLE SUMMARY:** **Strengths and limitations of this study**

- The study will serve as a large, randomised controlled trial providing grade A evidence on the most clinically- and cost-effective thromboprophylaxis regimen following superficial endovenous treatment.
- The primary outcome holds clinical significance.
- Using VTE prophylaxis may be associated with adverse clinical outcomes, increased risks and may not be cost-effective.
- Should pharmacological thromboprophylaxis be shown to offer no additional benefit to patients undergoing superficial endovenous intervention, stopping this practice has the potential to generate significant cost savings for healthcare providers.

## INTRODUCTION

Varicose veins, also known as superficial refluxing veins, affect up to 45% of the UK population (1). Varicose veins not only reduce physical and mental health-related quality of life, but also contribute significantly to chronic venous disease, which is responsible for over half of all cases of leg ulcers (2–4). Superficial endovenous treatment (SET), offers a minimally invasive approach that can conveniently be performed in an outpatient setting (5). Endovenous surgery stands as the recommended first-choice management for symptomatic varicose veins, in line with NICE guidelines [CG168] (5). This recommendation aligns with the European Society for Vascular Surgery Clinical Practice Guidelines, providing a level I recommendation for endovenous techniques (6). SET not only improves quality of life and facilitates venous ulcer healing, but also offers cost savings to healthcare providers (7–13). Annually, approximately 30,000 endovenous varicose vein procedures are carried out within the NHS, a number estimated to reach 68,800 with full adherence to NICE guidelines (14). Additionally, an estimated 30,000 to 40,000 procedures are undertaken annually in the private sector.

Despite its efficacy, SET is associated with thrombotic complications, presenting occurrences of venous thromboembolism (VTE) and endothermal heat-induced thrombosis (EHIT) at rates as high as 3.4%. VTE, encompassing deep vein thrombosis (DVT) and pulmonary embolism (PE), is a significant cause of disability and subsequent societal economic consequences (15). Hospital-acquired thrombosis (HAT), which is defined as the development of VTE within 90 days of a hospital episode, significantly contributes to morbidity and mortality, with statistics indicating a rate of 57 deaths per 100,000 admissions within the NHS (16). Complications following a DVT are substantial, with up to 50% of patients developing post thrombotic syndrome (PTS), characterised by chronic leg pain, oedema and skin changes (17,18). Additionally, PE is associated with lifelong functional and psychological repercussions, ultimately posing a risk of death during the index event (19,20). EHIT, encompassing any thrombus forming within 4 weeks of endovenous ablation and extending from the treated vein toward or into a deep vein, is categorised into four classes. EHIT classes 3 – 4 involve significant thrombus extension into and encompassing the deep vein, often necessitating treatment similar to that for DVT (9,10). SET presents a unique VTE risk compared to other short-stay surgical procedures, exhibiting a VTE rate comparable to that observed in major joint surgeries (16). Comparable day-case surgical procedures such as inguinal hernia repair or laparoscopic cholecystectomy exhibit a notably lower VTE rate of 0.3% (21).

In attempting to reduce the risk of VTE, UK clinicians exhibit varying approaches: 52% routinely prescribe a single dose of low-molecular weight heparin (LMWH) and 15% routinely prescribe extended pharmacological thromboprophylaxis with either LMWH or a direct-acting oral anticoagulant (DOAC), while 33% do not prescribe any pharmacological thromboprophylaxis (22). These practices align with findings from a 2019 national survey of vascular surgeons in Ireland, indicating that 73% of practitioners routinely prescribe pharmacological thromboprophylaxis for SET, utilising either a single dose of LMWH or extended prophylaxis, while 27% of practitioners do not prescribe any form of pharmacological thromboprophylaxis (23). Contradicting NICE guidelines, the routine use of pharmacological thromboprophylaxis for SET has become prevailing practice, despite having no supportive evidence base. A systematic review and meta-analysis published in 2022 failed to identify any high-quality evidence to support current pharmacological thromboprophylaxis strategies in this group of patients (24). However, the study suggested there is a significant reduction in the rate of DVT with additional pharmacological thromboprophylaxis (1.09% versus 3.20% for pharmacological thromboprophylaxis versus compression alone). International and national guidelines reflect the paucity of evidence in this area, with the European Society of Vascular Surgery guidelines providing a (IIa B) recommendation for consideration of individualised thromboprophylaxis strategies (6). This is exacerbated by NICE NG89 recommending that “prophylaxis is generally not needed for people undergoing varicose vein surgery” if their VTE risk assessment deems them low-risk (25).

Clinicians also lack confidence in current risk assessment tools (RATs) for patients undergoing SET (15). Despite the utilisation of RATs such as the Department of Health Risk Assessment (DHRA) Tool in the UK and the Caprini RAT in Europe and the US, none have undergone validation for varicose vein procedures. Consequently, varicose vein intervention-specific RATs have emerged, yet these, too, lack validation. Thus, there is currently no consensus on which risk factors provide clinical indication for pharmacological thromboprophylaxis or confer high-risk for VTE (26).

Pharmacological thromboprophylaxis presents potential clinical and cost benefits in VTE prevention, however grade A evidence is required to either support or refute this practice. Furthermore, as current RATs have not been validated in this patient group, unbiased prospective evidence will help guide risk-stratifying patients in the future. This research question also aligns with the James Lind Alliance priority setting for venous disease, further underlining the need for this trial.

### Objectives

The aims of this study are to establish whether in patients undergoing SET:

1. A single dose of pharmacological thromboprophylaxis decreases the risk of VTE
2. An extended course of pharmacological thromboprophylaxis decreases the risk of VTE
3. Pharmacological thromboprophylaxis is associated with an increased rate of bleeding events
4. Providing pharmacological prophylaxis is cost-effective
5. Any pharmacological thromboprophylaxis affects the rate of VTE

## METHODS AND ANALYSIS

### Trial design

This is a multi-centre, assessor-blind randomised controlled trial with a superiority comparison. The primary outcome is assessed blindly.

### Study setting

This trial will take place in NHS hospitals and private clinics delivering endovenous varicose vein procedures under local anaesthesia. Recruitment centres will need to have a pre-existing practice prior to the trial to prevent any learning curve effects.

### Eligibility criteria

Inclusion criteria are: adult patients (>18 years) scheduled to undergo endovenous intervention of truncal varicose veins under local anaesthesia. Treatment technologies include radiofrequency, laser, mechanochemical, foam sclerotherapy and cyanoacrylate glue.

Exclusion criteria are: clinical indication for therapeutic anticoagulation (e.g. atrial fibrillation, previous personal or first-degree relative history of VTE, thrombophilia). Also female patients of childbearing potential with a positive pregnancy test; those with a history of allergy to heparins or DOACs; a history of heparin-induced thrombocytopenia in the last year; inherited and acquired bleeding disorders; evidence of active bleeding; concomitant major health problems such as active cancer and chronic renal and/or liver impairment; known thrombocytopenia (platelets known to be less than 50 x 10^9^/l); surgery or major trauma in the previous 90 days; recent ischaemic stroke in the previous 90 days; inability to provide consent.

### Interventions

There are currently three thromboprophylaxis strategies used across the UK, with the trial arms of this application mirroring these practices (23). Intervention arms will consist of a single prophylactic dose of LMWH (e.g. dalteparin sodium, tinzaparin sodium, enoxaparin sodium) plus compression as per local practice (e.g. stockings, bandages, wraps, pads), and a single dose of LMWH plus extended thromboprophylaxis with LMWH or a DOAC (e.g. rivaroxaban, apixaban, dabigatran etexilate) plus compression. The choice between LMWH or DOAC for the extended thromboprophylaxis arm will be site-specific and dependent on local practice. The duration of this must be at least 7 days, but can be in line with local practice i.e., between 7 and 14 days in duration. The control arm will consist of compression (bandages, stockings, wraps or padding) as per local practice alone. The participant flow in the trial is displayed in *Figure 1*.

**Figure 1.**
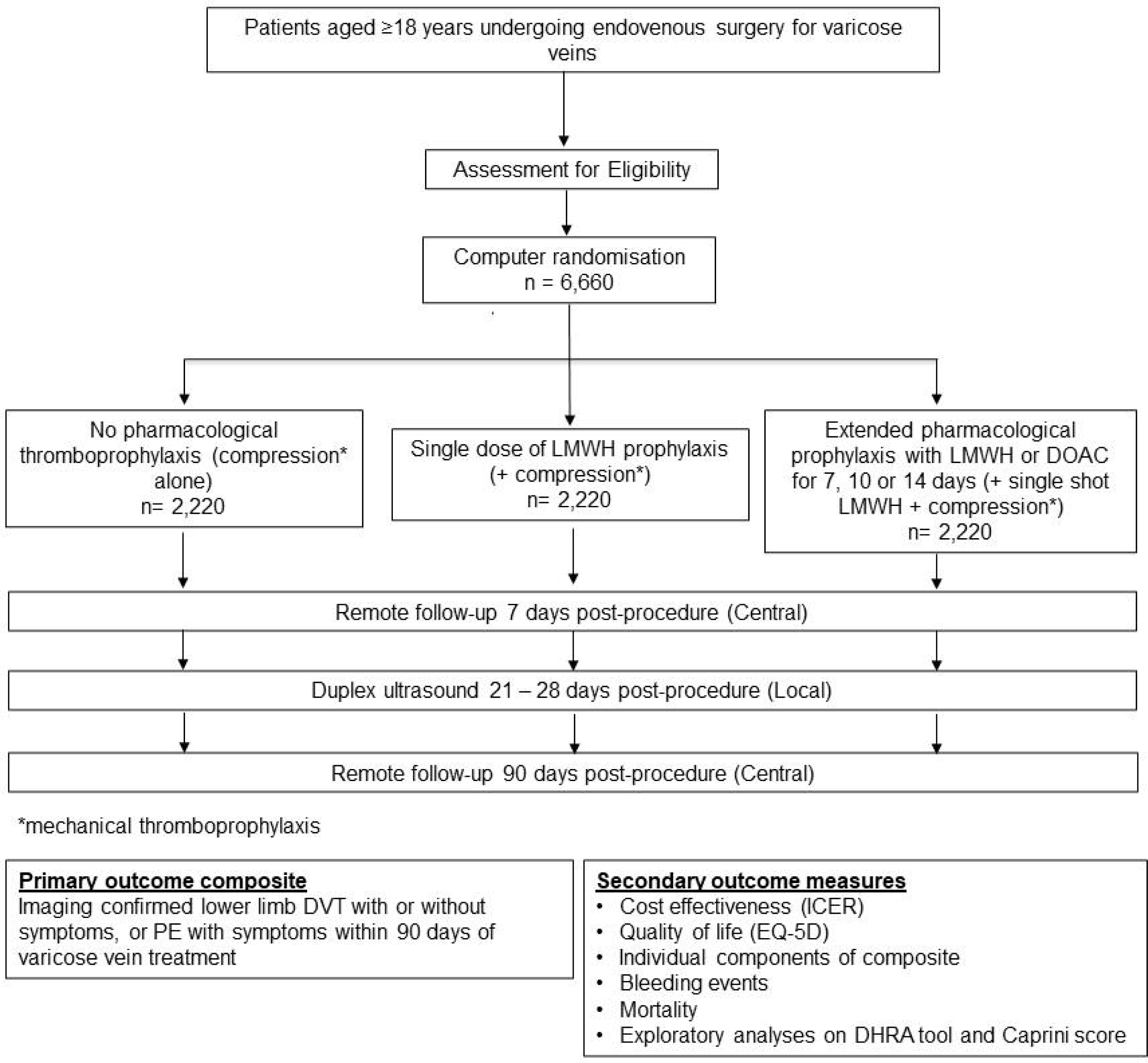
THRIVE study flow chart Abbreviations: DHRA, Department of Health Risk Assessment; DOAC, direct-acting oral anticoagulant; DVT, deep vein thrombosis; ICER, incremental cost-effectiveness ratio; LMWH, low-molecular weight heparin; VTE, venous thromboembolism.

### Primary outcome

Imaging confirmed lower limb DVT with or without symptoms, or PE with symptoms within 90 days of varicose vein treatment.

### Secondary outcomes

The secondary outcomes include:

1. Individual components of the composite outcome
2. Comparisons of quality of life at 7- and 90-days post-procedure using the EQ-5D
3. Mortality rates in each group
4. Cost-effectiveness of providing pharmacological thromboprophylaxis
5. Exploratory analyses to assess how well the DHRA tool and Caprini score predict outcome

### Safety outcomes

Safety monitoring includes any bleeding event. Major bleeding is defined as per the International Society on Thrombosis and Haemostasis standardised definition (27), which includes:

1. Bleeding into a critical organ
2. Bleeding into a surgical site requiring re-operation
3. Bleeding that leads to presentation to acute service

### Sample size and study duration

The most comprehensive evidence from 52 studies suggests that the rate of VTE (encompassing DVT, PE, EHIT) post-endovenous great saphenous vein interventions is 1.7% (1). However, this analysis did not investigate treatment effects. To address this, we conducted an analysis of 229 study arms, pooling data from 480,581 participants undergoing endovenous interventions. Our approach involved subgroup analyses, comparing those receiving pharmacological thromboprophylaxis and compression versus compression alone, and distinguishing between asymptomatic screen-detected and symptomatic VTE (24).

For individuals receiving pharmacological thromboprophylaxis (either single dose of LMWH or extended duration), rates of DVT, PE, and EHIT 3 - 4 were 0.521%, 0.216%, and 0.354%, respectively, resulting in a maximum summary VTE rate of 1.09%. Those receiving compression alone showed rates of 2.264% for DVT, 0.058% for PE, and 0.878% for EHIT 3-4, resulting in a maximum summary VTE rate of 3.20%. Rates of EHIT and DVT are distinct in this calculation, thus the impact of double counting contributing events will be negligible. In the subset data from RCT arms alone, rates of DVT were similar. When interpreting these figures in the context of the wider literature, balancing this with confounding by indication, and acknowledging that VTE rate will likely be lower in the compression alone arm due to the exclusion of individuals at the highest risk of VTE, we anticipate that the true value lies nearer to 1.0% in the pharmacological thromboprophylaxis arm and 2.7% in the compression alone arm.

At 90% power and 2.5% alpha (to approximately control overall alpha to 5%, with two active drugs being compared with a common control), the study could detect a significant change of 1.7% in 90-day VTE. This base case would require 1554 participants per group. Allowing for 10% crossover (which can only be control patients, compression only, receiving pharmacological thromboprophylaxis, since everyone receives compression) increases this to 1919 per group. Allowing for a single interim analysis at half time (50% randomised with 90 day follow up) for early stopping, for either futility or overwhelming evidence of efficacy analysis, inflates this to 1998 per group, under a two-sided, asymmetric, group sequential design implementing the non-binding Hwang-Shih-DeCani spending function [lower bound (futility) with gamma-2; upper bound (efficacy) with gamma= −4] (28). If we then allow 10% for loss to follow up, the total sample, randomised 1:1:1 between all groups, becomes 6660 (or 2220 per group).

We will more accurately estimate the required sample size by simulation in a sample size re-estimation step at around 20% mature data, using Dunnett’s 3-arm design with a common control, with correction, but for simplicity retaining the 1:1:1 equal randomisation; and inputting the observed missing data proportion at that stage.

### Interim analysis

There will be a formal interim analysis with the possibility of stopping early for futility (no prospect of a clinically meaningful treatment effect), at the point of 50% mature primary outcome data. Full details of the stopping boundaries and analysis will be detailed in a Statistical Analysis Plan (SAP), which the independent Data Monitoring Committee (iDMC) will approve prior to seeing unblinded data. The unblinded statistician will have no other role in the study while it is ongoing. The stopping rules are statistically non-binding. The iDMC may recommend early stopping of the study if the boundaries are crossed. They would make a recommendation to the independent Trial Steering Committee (TSC) who may or may not endorse that recommendation. The trial may stop at any time for safety if there is an excess of events in the intervention groups that is considered to generate avoidable harm. This decision would not be based on any statistical criterion and would be taken by the iDMC, then endorsed by the TSC.

### Recruitment and randomisation

Adults scheduled to undergo SET will be pre-screened by a member of the direct care team and invited to discuss the trial with a member of the research team. Informed consent will be obtained from each participant prior to participation in the trial, following adequate explanation of the aims, methods, anticipated benefits, and potential hazards of the trial. The individual obtaining consent will be a registered healthcare professional and will have been delegated this duty by the Principal Investigator (PI) on the delegation log. Participants may provide their consent to participate in the trial electronically or in written form. Participants will initial each statement on the consent form to confirm agreement. A copy of the consent form will be provided to the participant for their records. There is no defined timeframe between initial consent and the baseline visit, however confirmation that the participant’s consent is still valid prior to randomisation will be required in the baseline case report form (CRF).

Participants (n = 6,660) will undergo 1:1:1 web-based randomisation to one of three thromboprophylaxis strategies prior to undergoing SET. Randomisation will be conducted by an automated system linked to the Research Electronic Data Capture (REDCap) database setup via the Study Data Centre at the Edinburgh Clinical Trials Unit (ECTU), University of Edinburgh (a fully registered UKCRC Clinical Trials Unit, registration number 15). Recruitment will commence on 01 January 2024 for 27 months. The study will close on 31 December 2026.

### Blinding

Clinicians and participants will be aware of their treatment allocation. Assessors, being those who perform the venous duplex ultrasound scan and those responsible for collecting follow-up data at 7- and 90-days post-procedure, will be blinded to the treatment allocation.

### Follow-up periods

Participants will undergo a lower limb venous duplex ultrasound scan at 21 – 28 days post-intervention to identify asymptomatic DVT. This is timed to capture the peak onset of events which is at 3 weeks (29). Participants will be further followed up remotely by telephone, online or short message service (SMS) at 7- and 90-days with an expected VTE capture >95% (29). Longer term follow-up may be considered with award from a subsequent project grant to assess long-term efficacy of the intervention, hence contact at a later point will be included in the consenting process.

### Data collection and confidentiality

Participant data will be entered into the REDCap database by the local research teams. Source data stored at study centres will be archived locally as per local study operating procedures. Data and all appropriate documentation will be stored for a minimum of 10 years after the completion of the study, including the follow-up period. Details of procedures for CRF completion will be provided in a separate study manual. A formal Data Management Plan will be constructed to describe the procedures involved in the data management activities and processes for the study so that it is managed and maintained in accordance with the Good Clinical Practice (GCP) guidelines, local Research and Governance Integrity Team (RGIT) standard operating procedures, appropriate regulatory requirements, and the study protocol.

### Statistical analyses

All statistical analyses will be governed by a comprehensive SAP, written by the study statistician, and agreed by the TSC and iDMC before any unblinded data is seen. The main analysis will be according to the intention-to-treat principle where all consented participants will be included in the analysis retained in the group to which they were allocated (i.e., “as randomised”) and for whom outcome data are available. All results will be presented as point estimates, confidence intervals at the appropriate level and associated p-values. Absolute measures of effect will be presented alongside relative measures. The primary analysis will compare the incidence of VTE at 90 days, using a repeated measures analysis of variance (ANOVA), adjusting for any pre-specified strongly prognostic baseline covariates using a mixed effects logistic regression. Study site will be included in the model as a random effect and pre-specified baseline covariates strongly related to outcome will be included to adjust the estimated treatment effect. The findings will be assessed for robustness against any missing data, first using multiple imputation assuming this data is missing at random and, if appropriate and the data permits, further sensitivity analyses will be attempted under any plausible missing data mechanisms not missing at random. Secondary outcomes will be analysed in a similar fashion with generalised linear models appropriate to the distribution of the outcome. Safety data will be summarised descriptively.

### Internal pilot

There will be an internal pilot to assess feasibility of recruitment over nine months of recruitment to the trial, in which we will start recruiting from the (minimum) 40 centres. Site setup will be staggered over nine months, i.e., five centres per month. The target number of participants by the end of the 9-month internal pilot is 1450 participants. For the internal pilot, we will use stop-go criteria based on a Green-Amber-Red statistical approach (see *Table 1*).

**Table 1.**
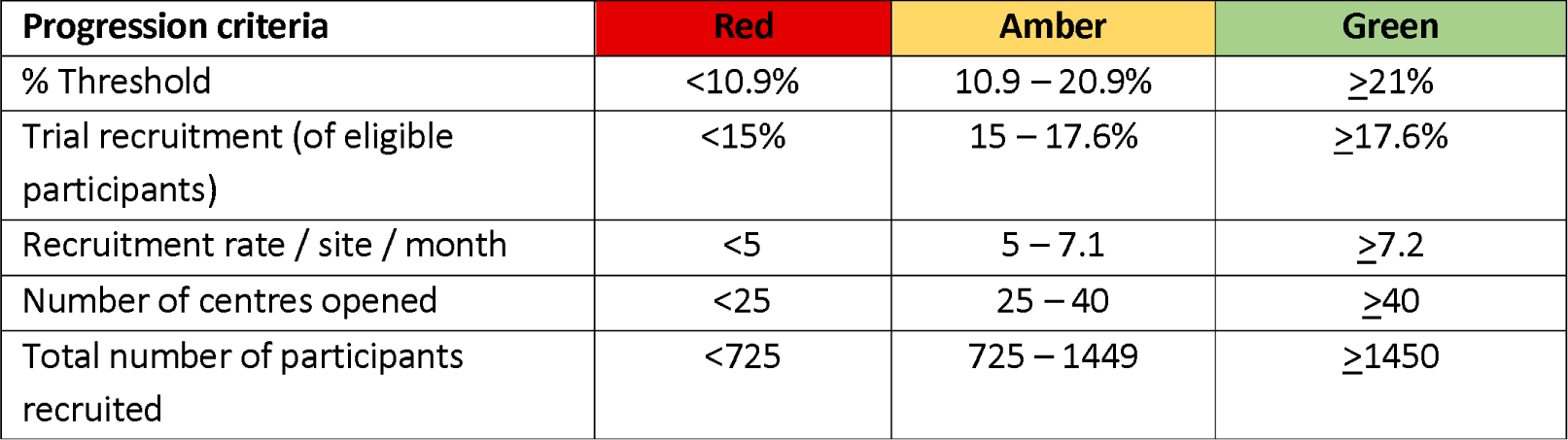
Internal pilot of feasibility assessment at 9 months.

| Progression criteria | Red | Amber | Green |
| --- | --- | --- | --- |
| % Threshold | <10.9% | 10.9 – 20.9% | ≥21% |
| Trial recruitment (of eligible participants) | <15% | 15 – 17.6% | ≥17.6% |
| Recruitment rate / site / month | <5 | 5 – 7.1 | ≥7.2 |
| Number of centres opened | <25 | 25 – 40 | ≥40 |
| Total number of participants recruited | <725 | 725 – 1449 | ≥1450 |

### Cost-effectiveness analysis

Two health economic analyses will be conducted, and a separate Health Economics Analysis Plan (HEAP) will be developed by the Health Economist detailing the proposed analyses. The main analyses will be performed from the perspective of the NHS and Personal Social Services, with secondary analyses from a societal perspective.

A within-trial analysis will compare the two pharmacological thromboprophylaxis strategies to compression alone over the 90 days of the study. Resource use items associated with treatments in primary and secondary care will be collected using case notes and self-completed patient diaries, and costed using manufacturer list prices, previous literature, and national reference costs. Days off work and normal activities and other patient-related costs will be collected for a secondary analysis. EQ-5D will be collected at baseline and follow up, analysed using the NICE approved tariff.

If the trial indicates that pharmacological thromboprophylaxis could be an effective therapy, a Markov (state–transition) decision model will be constructed to compare the cost effectiveness of the two pharmacological thromboprophylaxis strategies and compression alone over a longer time horizon. The time horizon of the model will be 2-years, allowing extrapolation of sequelae of VTE events (such as PTS) over the longer term to quantify the impact of VTE on patient health via quality adjusted life years (QALYs) and resource use. A preliminary model has been constructed based on published literature to identify the key variables that would need to be collected during the clinical study, and to estimate the number needed to treat (NNT) to avoid one VTE, above which pharmacological thromboprophylaxis would not be considered cost-effective at NICE thresholds. This model conservatively assumes 30% of patients with VTE develop PTS, with 3% of those having severe PTS.

The minimum cost of purchasing 10-days of thromboprophylaxis and providing allocated time for administration training equates to ∼£63.13 (30,31). Our model assumes cost of treatment of VTE, non-severe and severe PTS, as £451, £872 and £1,547 respectively and estimates of the respective utility decrement associated with symptomatic VTE and PTS are 0.8628, 0.7745 and 0.6752 respectively (32). Different durations of thromboprophylaxis will be modelled. These estimates will be reviewed at the time of the cost-effectiveness analysis, and any changes to these estimates will be updated.

Utilising a 2-year time horizon, incremental cost effectiveness ratio of pharmacological thromboprophylaxis in comparison to no pharmacological therapy would be £13,339 per QALY if the NNT were 59 participants (1/0.017). For pharmacological thromboprophylaxis to be cost effective at a NICE willingness to pay threshold of £20,000 per QALY, the NNT would need to be below 80.

Main analyses will be undertaken from perspective of the NHS and Personal Social Services, with secondary analyses from a societal perspective. Health economic analysis will be conducted according to NICE reference case and CHEERS guidelines, including sensitivity analyses and probabilistic sensitivity analyses (33,34). Results will be presented as estimates of mean incremental costs, effects, and incremental cost per QALY.

### Data monitoring, safety and quality control

An independent TSC and iDMC have been convened. The role of the TSC is to provide overall supervision of trial conduct and progress, and ensure the study adheres to GCP principles. The role of the iDMC is to oversee the safety of the trial participants, and the iDMC will be the only oversight committee that sees unblinded data as the trial progresses, which they will keep in strict confidence. Details of membership, responsibilities and frequency of meetings for the TSC and iDMC have been defined in separate Charters.

The Trial Management Group will oversee trial progress. All adverse events (AEs) will be collected and recorded on the REDCap database. Causality and relatedness will be assigned by the local PI or other appropriately trained and delegated individual. Research sites will inform the Coordinating Centre of all serious adverse events (SAEs) within 24 hours of knowledge of the event.

A study-specific risk assessment has been prepared in preparation for the study by the Trial Manager and study sponsor, which will be updated as required during the course of the trial. The frequency, type and intensity of monitoring visits will be detailed in a separate Data Monitoring Plan (DMP). The DMP will also detail the procedures for completion and sign-off of monitoring reports. In the event of a request for a trial site inspection by any regulatory authority, the study coordinating team must be notified as soon as possible. Participating investigators must agree to allow trial related monitoring, including audits, Research Ethics Committee (REC) review and regulatory inspections, by providing access to source data and other trial related documentation as required. Participant consent for this must be obtained as part of the informed consent process for the trial. The trial coordinating centre will centrally review eCRF data for errors and missing key data points on an ongoing basis.

Quality Control will be performed according to Imperial College internal procedures. The study may be audited by a Quality Assurance representative of the Sponsor. All necessary data and documents will be made available for inspection. The study may be subject to inspection and audit by regulatory bodies to ensure adherence to GCP and the UK Policy Framework for Health and Social Care Research.

### Patient and public involvement

Regular sustained patient and public involvement (PPI) has been undertaken to design and guide the study. Successful focus groups and interviews were held with patient representatives longitudinally throughout the trial development to gain insights into patients’ lived experiences and gather feedback on potential trial designs. Online surveys were also conducted with the wider venous community to inform the Plain English Summary. The patient-facing documentation has been designed in collaboration with three patient representatives. Two patient advisers have agreed to sit on the TSC, as well as review updated patient-facing documentation and assist with the dissemination of study results. An online survey of key stakeholders was also undertaken at the UK’s largest national conference to ascertain acceptability among clinicians of the proposed trial design.

## ETHICS AND DISSEMINATION

The Study Coordination Centre has obtained approval from the London Brent REC (23/LO/0261). Protocol amendments will be submitted to the sponsor for review before applying for approval from the REC, Health Research Authority (HRA) and Medicines and Healthcare Products Regulatory Agency (MHRA) and updating the ICRCTN record accordingly. The study must also receive confirmation of capacity and capability from each participating NHS Trust before accepting any research activity is carried out. The study will be conducted in accordance with the recommendations for physicians involved in research on human subjects adopted by the 18th World Medical Assembly, Helsinki 1964 and later revisions. Study findings will guide international clinical practice and stimulate key updates to international guidelines. Results will be published in high-impact journals alongside presentation at national and international vascular and haematology societies.

## Current study status

The study is currently in the process of setting up recruiting centres.

## Trial sponsor

Imperial College London is the main sponsor for this study. Delegated responsibilities are assigned to the NHS Trusts/Health Boards and non-NHS sites taking part in this study.

## Availability of data and materials

Data will be made available on reasonable request.

## Author contributions

The author contributions were as follows: Conception (AHD, DC and MM); Design of the work (AHD, DC, MM, SW, JN, LB, BJH, LB-S, JS, TE, MG, MW, SR, SO, BT, SN, RL, AS-B, CS and JD); Drafting the first version of the manuscript (SW); Critical review of the manuscript (AHD, DC, MM, JN, LB, BJH, LB-S, JS, TE, MG, MW, SR, SO, BT, SN, RL, AS-B, CS and JD).

## Data Availability

Data will be made available on reasonable request.

## Abbreviations

AE: Adverse event
ANOVA: Analysis of variance
CRF: Case report form
DHRA: Department of Health Risk Assessment
DOCA: Direct-acting oral anticoagulant
DVT: Deep vein thrombosis
ECTU: Edinburgh Clinical Trials Unit
EHIT: Endothermal heat-induced thrombosis
GCP: Good clinical practice
HAT: Hospital-acquired thrombosis
HEAP: Health economics analysis plan
HRA: Health Regulatory Authority
HTA: Health Technology Assessment
ICER: Incremental cost-effectiveness ratio
iDMC: Independent Data Monitoring Committee
LMWH: Low-molecular weight heparin
MHRA: Medicines and Healthcare Regulatory Agency
NHS: National Health Service
NICE: National Institute for Care and Excellence
NIHR: National Institute for Health Research
NNT: Number needed to treat
PE: Pulmonary embolism
PI: Principal Investigator
PPI: Patient and public involvement
PTS: Post-thrombotic syndrome
QALY: Quality adjusted life years
RAT: Risk assessment tool
RCT: Randomised controlled trial
REDCap: Research Electronic Data Capture
REC: Research Ethics Committee
SAE: Serious adverse event
SAP: Statistical analysis plan
SMS: Short message service
TSC: Trial Steering Committee
UK: United Kingdom
VTE: Venous thromboembolism

